# Diagnostic Performances of GPT-4o, Claude 3 Opus, and Gemini 1.5 Pro in “Diagnosis Please” Cases

**DOI:** 10.1101/2024.05.26.24307915

**Authors:** Yuki Sonoda, Ryo Kurokawa, Yuta Nakamura, Jun Kanzawa, Mariko Kurokawa, Yuji Ohizumi, Wataru Gonoi, Osamu Abe

## Abstract

**Backgrounds:** Large language models (LLMs) are rapidly advancing and demonstrating high performance in understanding textual information, suggesting potential applications in interpreting patient histories and documented imaging findings. LLMs are advancing rapidly and an improvement in their diagnostic ability is expected. Furthermore, there has been a lack of comprehensive comparisons between LLMs from various manufacturers.

**Purpose:** We tested the diagnostic performance of the latest three major LLMs (GPT-4o, Claude 3 Opus, and Gemini 1.5 Pro) using *Radiology* Diagnosis Please cases, a monthly diagnostic quiz series for radiology experts.

**Materials and Methods:** Clinical history and imaging findings as provided textually by the case submitters were extracted from 324 quiz questions from *Radiology* Diagnosis Please cases. The GPT-4o, Claude 3 Opus, and Gemini 1.5 Pro generated the top three differential diagnoses. Diagnostic performance among the three LLMs were compared using Cochrane’s Q and post-hoc McNemar’s tests.

**Results:** The diagnostic accuracies for the primary diagnosis were 41.0%, 54.0%, and 33.9% for GPT-4o, Claude 3 Opus, and Gemini 1.5 Pro, respectively. When considering the accuracy of any of the top three differential diagnoses, the rates improved to 49.4%, 62.0%, and 41.0%, respectively. Significant differences in diagnostic performance were observed among all pairs of the models.

**Conclusion:** In a comparison of the latest LLMs, Claude 3 Opus outperformed GPT-4o and Gemini 1.5 Pro in solving radiology quiz cases. These models appear capable of assisting radiologists when supplied with accurate evaluations and worded descriptions of imaging findings by radiologists.

**Summary statement:** Claude 3 Opus achieved the highest diagnostic accuracy, followed by GPT-4o and Gemini 1.5 Pro, in a comparison of their performance on 324 text-based *Radiology* Diagnosis Please cases..

**Key Results:**

- This study compared the diagnostic performances of the latest three major large language models, GPT-4o, Claude 3 Opus, and Gemini 1.5 Pro, using clinical history and textualized imaging findings in *Radiology* Diagnosis Please cases.
- The top three differential diagnoses generated by GPT-4o, Claude 3 Opus, and Gemini 1.5 Pro achieved diagnostic accuracies of 49.4%, 62.0%, and 41.0%, respectively, with statistically significant differences between each model’s performance.

## Introduction

Large language models (LLMs) are neural network models trained on vast amounts of text data, which demonstrate high performance in natural language processing tasks(1). They can be utilized across different fields, including medicine(2).

Previous research has investigated the diagnostic capabilities of LLMs in radiology. In a study by Ueda et al.(3), OpenAI’s GPT-4 model(4) correctly answered 54% (170/313) of Diagnosis Please cases, a monthly diagnostic radiology quiz case series for radiology experts published in an international academic journal, *Radiology*, relying solely on the provided clinical history and imaging findings. Similarly, Horiuchi et al.(5) reported that the GPT-4 model achieved a diagnostic accuracy of 50% (50/100 cases) in “Case of the Week’’ diagnostic quiz case series published in a academic journal American Journal of Neuroradiology, also based on the given clinical history and imaging findings.

Recently, various manufacturers have developed their own LLMs, which have undergone multiple version upgrades. Flagship models include OpenAI’s GPT-4o model(6), Anthropic’s Claude 3 Opus(7), and Google’s Gemini 1.5 Pro(8). Li et al.(9) demonstrated a significant improvement in diagnostic performance using the GPT-4 model compared with the GPT-3.5 Turbo model. This finding suggests that version upgrades can lead to improvements in the diagnostic performances of LLMs.

In a previous study, we evaluated the diagnostic performance of Claude 3 Opus using Diagnosis Please cases and reported a diagnostic accuracy of 62.1% for top three differential diagnoses using textual clinical history and imaging findings(10). However, the diagnostic abilities of the recently released GPT-4o and Gemini 1.5 Pro have not yet been investigated.

Therefore, in this study, we assessed and compared the diagnostic performance of three flagship models - GPT-4o, Claude 3 Opus, and Gemini 1.5 Pro - on Diagnosis Please cases using clinical history and imaging findings. This research aims to provide insights into the current state-of-the-art in LLM-based diagnostic capabilities and highlight the potential differences between these advanced models.

## Materials and Methods

We used GPT-4o (OpenAI, San Francisco, United States, released on May 13, 2024), Claude 3 Opus (Anthropic, California, United States, released on March 4, 2024), and Gemini 1.5 Pro (Google, Mountain View, United States, released on April 9, 2024) to list a primary diagnosis and two differential diagnoses for 324 quiz questions (cases 1 to 324, published between 1998 and 2023) from *Radiology* Diagnosis Please (https://dxp.rsna.org/).

We used application programming interfaces to access each model (GPT-4o: gpt-4o-2024-05-13, Claude 3 Opus: claude-3-opus-20240229, Gemini 1.5 Pro: gemini-1.5-pro-latest, accessed on May 18, 2024). For all models, we specified the generation parameters as temperature=0.0 and top-p=1.0. To prevent previous inputs from influencing subsequent ones, we conducted the input for each case on an independent session. The prompt was as follows(5,11): “*As a physician, I plan to utilize you for research purposes. Assuming you are a hypothetical physician, please walk me through the process from differential diagnosis to the most likely diagnosis and the next two most likely differential diagnoses step by step, based on the attached patient’s information*.”

When extracting the submitter-identified imaging findings, one trainee radiologist and one board-certified diagnostic radiologist with 11 years of experience meticulously removed sentences containing answers to ensure analysis integrity. The accuracy of the LLMs’ primary diagnosis and two differential diagnoses was determined by a consensus of three board-certified diagnostic radiologists with 8, 11, and 19 years of experience. Ambiguous answers or those lacking sufficient elements were classified as incorrect.

As this study utilized published articles, no ethical approval was required.

Cochrane’s Q test was used to assess the difference in performance between the three LLMs. When significant differences were detected, post hoc analyses were conducted using McNemar’s tests with continuity correction and Bonferroni’s correction to evaluate the differences in the accuracy rates for the top three differential diagnoses between each pair of models. Two-sided P-values < 0.05 were considered statistically significant. Statistical analyses were performed using R (version 4.1.1; R Foundation for Statistical Computing, Vienna, Austria).

## Results

The diagnostic accuracies for the primary diagnosis were 41.0%, 54.0%, and 33.9% for GPT-4o, Claude 3 Opus, and Gemini 1.5 Pro, respectively. The accuracy rates increased to 49.4%, 62.0%, and 41.0% for GPT-4o, Claude 3 Opus, and Gemini 1.5 Pro, respectively, when considering any of the top three differential diagnoses. Notably, for 6 out of 324 questions, Gemini 1.5 Pro responded with “Providing a differential diagnosis based on the information provided would be irresponsible and potentially harmful”, which were judged as incorrect answers.

Cochrane’s Q test demonstrated significant differences in diagnostic performance among the three LLMs (p < 0.001). Post hoc pairwise comparisons using McNemar’s tests with continuity correction and Bonferroni’s correction revealed that Claude 3 Opus outperformed GPT-4o (p < 0.001), which in turn outperformed Gemini 1.5 Pro (p = 0.001). Significant differences were observed between all combinations of the LLMs (Table 1).

**Table 1.** Diagnostic accuracy of each model, Cochrane’s Q test, and McNemar’s tests between each model.

|  | Accuracy [%] |  |  | Cochrane's Q test | McNemar's test |  |  |
| --- | --- | --- | --- | --- | --- | --- | --- |
|  | GPT-4o | Claude 3 Opus | Gemini 1.5 Pro |  | GPT-4o vs Claude 3 Opus | GPT-4o vs Gemini 1.5 Pro | Claude 3 Opus vs Gemini 1.5 Pro |
| Primary diagnosis | 41.0 (133/324) | 54.0 (175/324) | 33.9 (109/324) |  |  |  |  |
| Top 3 differential diagnoses | 49.4 (160/324) | 62.0 (201/324) | 41.0 (132/324) | p<0.001 | p<0.001 | p=0.001 | p<0.001 |

## Discussion

In our study, we compared the diagnostic performance of flagship LLMs from three companies on *Radiology* Diagnosis Please cases based on the provided clinical history and imaging findings. The results showed that the models performed in the following order from best to worst: Claude 3 Opus, GPT-4o, and Gemini 1.5 Pro. The differences between all pairwise combinations were statistically significant.

It is worth noting that as of the time of writing, the technical report for GPT-4o has not been released. However, Claude 3 Opus has reportedly outperformed Gemini 1.5 Pro in eight text-based language benchmarks, including reasoning, coding, and mathematics(7).

In the context of medical natural language processing capabilities, despite being a general-purpose LLM, Claude 3 Opus achieved an accuracy of 74.9% for 0-shot and 75.8% for 5-shot on PubMedQA(12), which is nearly equivalent to the performance of Google’s Med-PaLM 2(13), an LLM specialized in medicine.

Regarding Gemini 1.5 Pro, one of its design philosophies is the extension of context length(8). The developers have also released Gemini 1.5 Flash, a lightweight and fast model with slightly reduced performance(14). These points suggest that Gemini 1.5 series may prioritize real-world implementations, such as integration into devices, over benchmark performance.

In this study, the accuracy of GPT-4o is lower than the previously reported results for GPT-4(3). One possible reason for this discrepancy is the strict grading criteria employed in our research. This issue arises from the fact that the actual correct answer criteria for *Radiology* Diagnosis Please cases are not publicly available, which is one of the limitations of this study.

In a previous study where GPT-4 Turbo with Vision were tasked with solving the Japanese Board of Radiology examination(10), GPT-4 Turbo with Vision, given both image and textual information, could not outperform GPT-4 Turbo which was only provided with textual information. Even Claude 3 Opus, which achieved the best performance in this study, showed significantly inferior diagnostic performance when given only the history and key images as input, without the textual information of imaging findings, compared to when the textual information of both history and imaging findings were provided, as we reported in our previous research(10). Therefore, at least at present, the main role of LLMs is not to replace radiologists but rather to assist in diagnosis using imaging findings based on accurate interpretations and verbalization of imaging findings by radiologists. However, to utilize the rapidly evolving LLMs in the field of diagnostic radiology, it is desirable to continue conducting research and evaluations in the future.

## Data Availability

All data produced are available online at https://pubs.rsna.org/journal/radiology.

## Abbreviations

AI: artificial intelligence
LLM: large language model

